# Clinical signatures of *SYNGAP1-*related disorders through data integration

**DOI:** 10.1101/2024.10.02.24314452

**Authors:** Jillian L. McKee, Jan H. Magielski, Julie Xian, Stacey Cohen, Jonathan Toib, Chen Chen, Dan Kim, Aakash Rathod, Elise Brimble, Nasha Fitter, J. Michael Graglia, Kathryn A. Helde, Michael J. Boland, Sarah McKeown Ruggiero, Rob Sederman, Ingo Helbig

**Affiliations:** Division of Neurology, Children’s Hospital of Philadelphia, Philadelphia, PA 19104, USA; The Epilepsy NeuroGenetics Initiative (ENGIN), Children’s Hospital of Philadelphia, Philadelphia, PA 19104, USA; Department of Biomedical and Health Informatics (DBHi), Children’s Hospital of Philadelphia, Philadelphia, PA 19146, USA; Epilepsy and Neurodevelopmental Disorders Center (ENDD), Children’s Hospital of Philadelphia, University of Pennsylvania Perelman School of Medicine, Philadelphia, PA 19104, USA; Ambit RD, Inc; Citizen Health, San Francisco, CA 94112, USA; Department of Physiology, University of Pennsylvania Perelman School of Medicine, Philadelphia, PA 19104, USA; Department of Neurology, University of Pennsylvania Perelman School of Medicine, Philadelphia, PA 19104, USA; SynGAP Research Fund, San Diego, CA

**Author notes:** Corresponding author: Ingo Helbig, MD; Corresponding author’s address: Division of Neurology, Children’s Hospital of Philadelphia, Philadelphia, PA 19104; Corresponding author’s; Corresponding author’s.

## Abstract

**Objective:** To characterize the longitudinal disease and treatment histories of individuals with *SYNGAP1-*related disorders.

**Methods:** Participants with *SYNGAP1* were identified from a range data data sources, including insurance claims data by ICD-10 diagnosis codes (*n*=246), a specialized medical record registry and a local cohort followed at a single tertiatry health care institution (*n*=158).

**Results:** Compared to a broader population of individuals with epilepsy, phenotypes associated with *SYNGAP1* disorders included behavioral abnormalities (Odds ratio (OR) 12.35, 95% CI 9.21–16.78), generalized-onset seizures (OR 1.56, CI 1.20–2.02), and autism (OR 12.23, CI 9.29–16.24). A wide range of clinical features showed distinct age-related patterns, such as a more than five-fold risk of autistic behavior emerging between 27 and 30 months. Generalized-onset seizures became significantly enriched (OR 4.05, CI 2.02–7.59) after 3 years of age and persisted over time. Valproic acid (OR 2.26, CI 1.29–3.70) and clobazam (OR 2.58, CI 1.55–4.09) were commonly used for epilepsy management, which contrasted significantly from treatment strategies in the broader epilepsy cohort. Furthermore, valproate and lamotrigine were more effective at reducing seizure frequencies or maintaining seizure freedom than other anti-seizure medications. Risperidone, aripiprazole, and guanfacine were commonly used for behavioral features.

**Interpretation:** Phenotypic features specific to *SYNGAP1* included a predominance and age-dependence of generalized seizures, a more than ten-fold risk of behavioral abnormalities, and a developmental profile with prominent deficits in verbal skill acquisition. Clear delineation of trajectories of *SYNGAP1-*related disorders will improve diagnosis, prognosis, and clinical care, facilitating clinical trial readiness.

## INTRODUCTION

Disease-causing variants in *SYNGAP1* are among the most common monogenic etiologies for generalized epilepsy and non-syndromic intellectual disability, with an estimated prevalence of 6 per 100,000.^1^ Since its first description in human epilepsy in 2009,^2^ pathogenic variants *SYNGAP1* are now recognized as a common cause of developmental and epileptic encephalopathies (DEEs) in addition to several distinct epilepsy syndromes, such as epilepsy with myoclonic atonic seizures (EMAtS) and epilepsy with eyelid myoclonia (EEM).^3–8^ However, the full range of clinical presentations in individuals with *SYNGAP1-*related disorders (*SYNGAP1*-RD) and the trajectories of symptoms over time has only been insufficiently delineated. This knowledge gap represents a major impediment for precision medicine trials as the longitudinal course of *SYNGAP1*-RD is incompletely understood.

*SYNGAP1* encodes a key regulatory protein of the post-synaptic density (PSD) at the interface of NMDA receptors and the downstream signaling apparatus, which is essential for synaptic plasticity.^9–11^ The main disease mechanism is haploinsufficiency, and the majority (∼74%) of individuals with *SYNGAP1*-RD have protein-truncating variants.^12^ *SYNGAP1* represents a focused target for novel precision medicine approaches including the development of antisense oligonucleotides (ASOs).^13–15^

Clinical research on *SYNGAP1*-RD has largely been performed through small cohort studies with focused phenotypic descriptions, and only limited genotype-phenotype associations have been described to date.^6,7,12^ Furthermore, especially in early childhood prior to epilepsy onset in *SYNGAP1-*RD, the phenotypic picture is frequently non-specific and the presentation of individuals with *SYNGAP1*-RD may resemble other neurodevelopmental conditions such as Rett Syndrome and Angelman Syndrome.^16,17^ An early molecular diagnosis has implications in the clinical management, including early intervention and tailored care for improving long-term developmental outcomes. Accordingly, early symptom recognition and clear delineation of longitudinal trajectories in *SYNGAP1*-RD remain critical.

Here, we characterize phenotypic features, including behavioral abnormalities and seizure types, enriched in individuals with *SYNGAP1*-RD compared with the broader population of individuals with epilepsy and neurodevelopmental disorders across the age span, outlining the longitudinal landscape of epilepsy and developmental trajectories using real-world data captured from various large-scale healthcare resources.

## METHODS

### Inclusion of individuals with SYNGAP1-RD from various healthcare resources

We identified individuals with *SYNGAP1-*RD through large-scale healthcare claims data using the International Classification of Diseases,^18^ Tenth Revision, Clinical Modification (ICD-10-CM) diagnosis code for *SYNGAP1* (F78.A1). Inclusion criteria were: (1) *SYNGAP1* F78.A1 diagnosis codes across at least two distinct encounters, or (2) an epilepsy, developmental delay, or intellectual disability diagnosis in addition to a *SYNGAP1* F78.A1 diagnoses. Only individuals under the age of 25 years were included, as the recency of the ICD10-code limited duration of clinical histories captured in the database for older individuals. We then included all individuals in the Citizen Health Natural History Registry and individuals with *SYNGAP1-*RD who received care at Children’s Hospital of Philadelphia (CHOP). To account for cohort overlap, we analyzed healthcare claims data separately from electronic medical record data from Citizen and our institution. Individuals with data in both the Citizen and CHOP datasets were identified by common variant, sex, and age, and any duplicates were removed.

### Longitudinal phenotypic analysis of the clinical landscape

First, we analyzed *SYNGAP1-*RD through healthcare claims data (*n*=246), which enabled us to analyze clinical histories including clinical diagnoses, phenotypic features, and treatment patterns. For treatment strategies, we assessed the current medication landscape for anti-seizure medications (ASMs) in addition to medications for behavioral features including anxiety, aggression, and sleep-related disorders.

Next, we assessed clinical histories in a smaller cohort (*n*=158) focusing on epilepsy trajectories and developmental outcomes from reconstructed medical records. We used a published framework championed by the Epilepsy Learning Health System (ELHS)^19^ and Pediatric Epilepsy Learning Health System (PELHS)^20^ and used previously by our group^21,22^ to capture seizure severity on a monthly basis, in which seizure frequencies (SF) are indicated by: multiple daily seizures (>5 per day, SF score = 5), several daily seizures (2–5 per day, SF score = 4), daily seizures (SF score = 3), weekly seizures (SF score = 2), monthly seizures (SF score = 1), and no seizures (SF score = 0). To assess development, we analyzed milestone acquisition. Clinical features and seizure types were captured using the Human Phenotype Ontology (HPO), a framework for phenotypic data harmonization.^23–26^ Clinical diagnoses were mapped to the HPO using the Unified Medical Language System (UMLS) crosswalk,^27^ supplemented by manual curation of seizure-related codes.

### Comparative ASM effectiveness analysis

Following the retrieval of ASM prescription data and the monthly reconstruction of seizure frequencies, we performed a comparative ASM effectiveness in individuals with *SYNGAP1-*RD from the Citizen Health dataset or who were seen at CHOP, as described previously.^21,22,28^ A total of 94 individuals had ASM prescription and seizure frequency information available.

To determine the comparative effectiveness of ASMs in individuals with *SYNGAP1*-RD, we analyzed how the SF scores were changing when they were prescribed different medications. For example, if an individual was treated with levetiracetam between two and four years of age, changes in SF scores at that time were compared to SF scores when they were not treated with levetiracetam. Fisher’s exact test was performed to assess how different ASMs are comparatively effective in (1) reducing seizure frequency and (2) maintaining seizure freedom. Only medications that were prescribed to at least 5 individuals were considered in the comparative ASM effectiveness analysis.

## RESULTS

### Individuals with SYNGAP1-RD can be identified through various healthcare resources

We identified individuals with *SYNGAP1*-RD through a combination of healthcare claims data, medical record aggregators, and individuals seen at a single tertiary pediatric healthcare center. 246 individuals with *SYNGAP1-*RD through healthcare claims data, spanning 1,321 cumulative patient-years. Reconstructed medical records of 158 individuals were included from the Citizen Health Natural History Registry (*n*=138) and Children’s Hospital of Philadelphia (*n*=20) across a total of 1,253 cumulative patient-years. The median age of inclusion was 6.53 years (IQR 1.5 – 12.5 years) and 5.6 years (IQR 3.8 – 10.0 years) in each respective cohort, and the median observation time was 6 years (IQR 4.8 – 6.3 years) and 5.4 years per individual (IQR 3.8 – 7.9 years) in the claims data and medical record cohorts, respectively. The median age of genetic diagnosis was 3.8 years (*n*=148 individuals where genetic testing was available, IQR 2.7 – 6.8 years). In the claims dataset, age at diagnosis was not available, but the median age at first *SYNGAP1* diagnosis code was 11.2 years (IQR 5.6 – 16.5 years).

The demographic and clinical characteristics across both cohorts were similar, but with a few notable differences (**Table 1**). For example, 131 of 158 individuals (82.9%) from reconstructed medical records had seizures, compared to 161 of 245 (65.4%) individuals identified through claims data with *SYNGAP1*-RD. Claims data also had relatively lower documented rates of neurodevelopmental delay, autism, and/or intellectual disability (78.9% compared to 100%) and behavioral features (75.2% compared to 95.7%).

**Table 1.** Cohort of individuals with *SYNGAP1*-RD.

|  | Healthcare claims | Electronic medical records |
| --- | --- | --- |
| **Cohorts (ICD10-CM)<sup>1</sup>** |  |  |
| *SYNGAP1*-related disorders (F78A1) | *n*=246 individuals | *n*=158 individuals |
| Broad epilepsy cohort (G40+) | *n*=680,076 individuals | -- |
| Angelman Syndrome (Q9351) | *n*=3,697 individuals | -- |
| Rett Syndrome (F842) | *n*=5,489 individuals | -- |
| **Demographics of *SYNGAP1* cohort** |  |  |
| Male, female | 140 male, 106 female | 80 male, 78 female |
| Median age at assessment (IQR) | 6.53 years (1.5 – 12.5 years) | 5.6 years (3.8 – 10.0 years) |
| Median observation time (IQR) | 6 years (4.8 – 6.3 years) | 5.4 years (3.8 – 7.9 years) |
| Median age of diagnosis (IQR) | 11.2 years (5.6 – 16.5 years)<sup>2</sup> | 3.8 years (*n*=148, 2.7 – 6.8 years) |
| Total patient-years | 1,321 years | 1,253 years |
| Variant spectrum | -- | 133 PTV/del<sup>3</sup>, 21 missense, 2 complex indels |
| **Clinical characteristics** |  |  |
| Epilepsy | 65.4% (*n*=161/246) | 82.9% (*n*=131/158) |
| Median age at seizure onset (IQR) | -- | 2.8 years (2.0 – 3.8 years) |
| Generalized-onset seizures | 66.5% (*n*=107/161) | 69.5% (*n*=91/131) |
| Bilateral tonic-clonic seizures | 46.6% (*n*=75/161) | 16.8% (*n*=22/131) |
| Atonic seizures | 32.9% (*n*=53/161) | 42.0% (*n*=55/131) |
| Absence seizures | 18.0% (*n*=29/161) | 54.2% (*n*=71/131) |
| Myoclonic seizures | 0.62% (*n*=1/161) | 22.9% (*n*=20/131) |
| NDD/autism/intellectual disability | 78.9% (*n*=194/246) | 100% (*n*=138/138) |
| Behavioral features | 75.2% (*n*=185/246) | 95.7% (*n*=132/138) |
| Sleep disturbances | 37.3% (*n*=69/185) | 74.6% (*n*=103/138) |
| Anxiety-related disorders | 11.9% (*n*=22/185) | 23.9% (*n*=33/138) |
| Aggressive behavior | 5.9% (*n*=11/185) | 52.2% (*n*=72/138) |

<sup>1</sup> We included individuals 25 years of age or younger for all cohorts: *SYNGAP1*-related disorders, Broad epilepsy cohort, Angelman Syndrome, and Rett Syndrome.
<sup>2</sup> Indicated by first clinical diagnosis using International Classification of Diseases, Tenth Revision, Clinical Modification (ICD-10-CM) diagnosis code for *SYNGAP1* (F78A1), as reflected in claims data.
<sup>3</sup> Protein truncating variant and deletions (PTV/del) included nonsense variants, frameshift variants, splice site variants, and whole or partial gene deletions.

### Phenotypic footprints of SYNGAP1 are distinct from other neurodevelopmental disorders

In order to assess disease-specific signatures or phenotypic footprints related to *SYNGAP1*-RD, we defined a broader epilepsy cohort of 680,076 individuals with a G40+ diagnosis and assessed cumulative diagnoses of G40+ alongside *SYNGAP1* (**Fig S1**). When comparing individuals with *SYNGAP1* with the broader epilepsy cohort, we found 95 significant associations after correcting for multiple testing (**Fig 1A**). Of note, individuals with *SYNGAP1-*RD were more likely to have behavioral abnormalities (Odds ratio (OR) 12.35, 95% CI 9.21 – 16.78), generalized-onset seizures (OR 1.56, 95% CI 1.20 – 2.02), autism (OR 12.23, 95% CI 9.29 – 16.24), and abnormality of higher mental function (including intellectual disability, OR 6.38, 95% CI 4.89 – 8.37). In contrast, individuals with *SYNGAP1* were less likely to have motor seizures than those in the broader G40+ cohort (OR 0.273, 95% CI 0.212 – 0.353).

**Figure 1.**
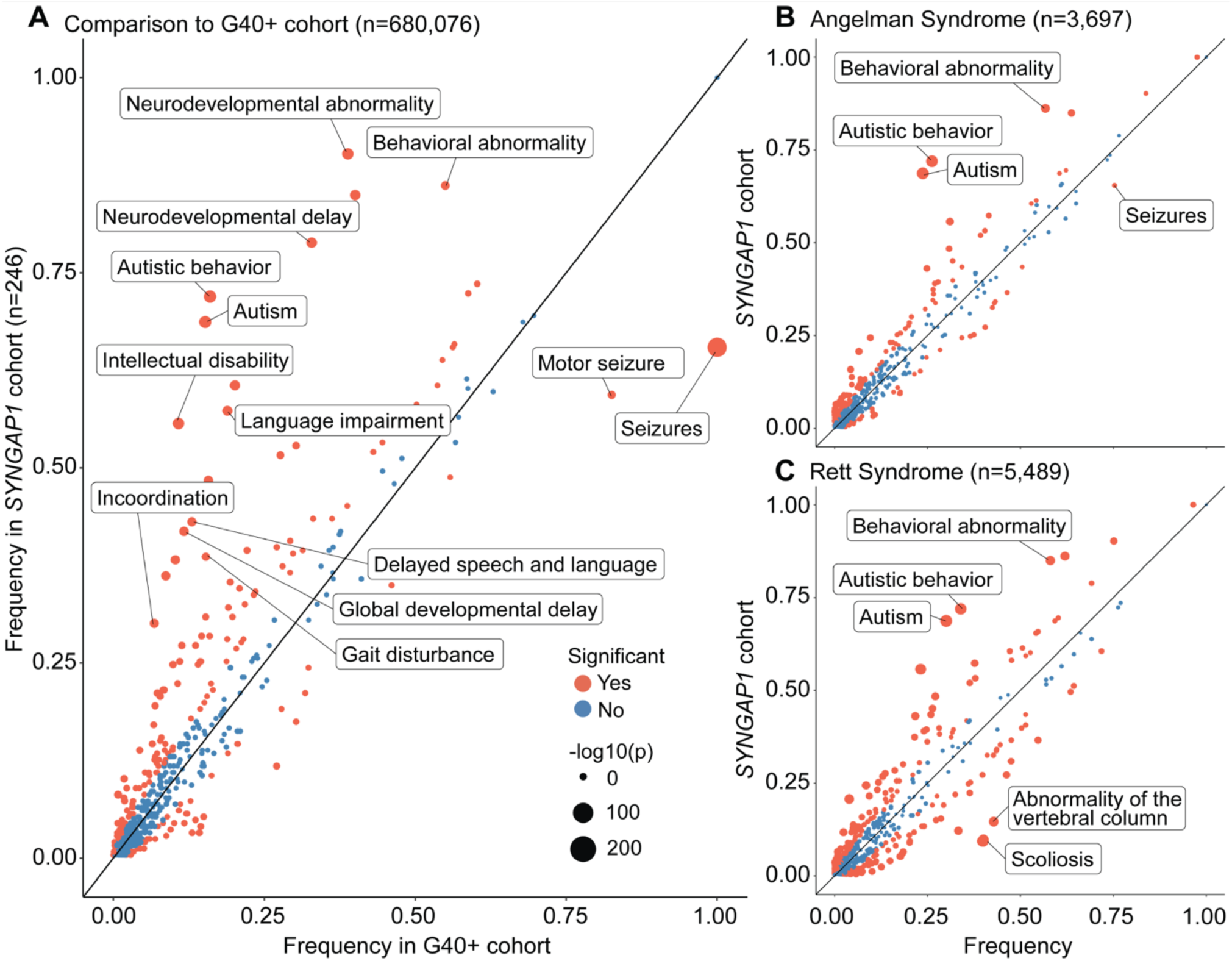
Clinical phenotypes enriched in individuals with *SYNGAP1* in comparison to other epilepsies and neurodevelopmental disorders. Individuals with *SYNGAP1-*RD were compared to a broader epilepsy cohort of 680,076 individuals with an epilepsy (G40+) diagnosis (A), as well as with syndromic comparator groups that phenotypically resembled *SYNGAP1*: 3,697 individuals with Angelman Syndrome (B) and 5,489 individuals with Rett Syndrome (C). Compared to the broad epilepsy cohort, behavioral abnormalities (Odds ratio (OR) 12.35, 95% CI 9.21 – 16.78), generalized-onset seizures (OR 1.56, 95% CI 1.20 – 2.02), autism (OR 12.23, 95% CI 9.29 – 16.24), and abnormality of higher mental function (including intellectual disability, OR 6.38, 95% CI 4.89 – 8.37) were enriched in individuals with *SYNGAP1*. When comparing *SYNGAP1* with Angelman and Rett syndromes, behavioral features (AS OR 7.2, 95% CI 5.3 – 9.9, RS OR 5.0, 95% CI 3.7 – 6.8) and autism (AS OR 6.8, 95% CI 5.1 – 9.2; RS OR 4.4, 95% CI 3.3 – 6.8) were more common.

We then contrasted individuals with *SYNGAP1-RD* with comparator groups of individuals that phenotypically resembled *SYNGAP1*, particularly during early development, including (1) 3,697 individuals with Angelman Syndrome and (2) 5,489 individuals with Rett Syndrome (**Fig 1B** and **1C**). When comparing *SYNGAP1* with Angelman (AS) and Rett (RS) syndromes, behavioral features remained enriched in those with *SYNGAP1* (AS OR 7.2, 95% CI 5.3 – 9.9, RS OR 5.0, 95% CI 3.7 – 6.8), as did autism (AS OR 6.8, 95% CI 5.1 – 9.2; RS OR 4.4, 95% CI 3.3 – 6.8). Additional clinical features included higher rates of abnormal verbal communicative behavior (AS OR 3.4, 95% CI 1.8 – 6.2, RS OR 2.4, 95% CI 1.3 – 4.2), increased typical absence seizures (AS OR 2.6, 95% CI 1.7 – 4.0, RS OR 5.3, 95% CI 3.3 – 8.1), and decreased relative frequency of status epilepticus (AS OR 0.68, 95% CI 0.48 – 0.95, RS OR 0.64, 95% CI 0.45 – 0.89).

### Longitudinal trajectories demonstrate age-specific phenotypic patterns, including a later seizure onset and ongoing seizures in SYNGAP1

We then assessed clinical features across the age span (**Fig 2**). We found that the overall clinical presentation of *SYNGAP1-* RD begins to diverge from comparator groups by the second year of life. When compared to the general epilepsy cohort, we found that individuals with *SYNGAP1*-RD were more likely to have behavioral abnormalities, first significant between 27 and 30 months (OR 3.00, 95% CI 1.50 – 5.68) and persisting throughout the lifespan. Autistic behavior also became prominent between 27 and 30 months (OR 5.71, 95% CI 2.44 – 11.9). Generalized-onset seizures became significantly enriched (OR 4.05, 95% CI 2.02 – 7.59) after 3 years of age.

**Figure 2.**
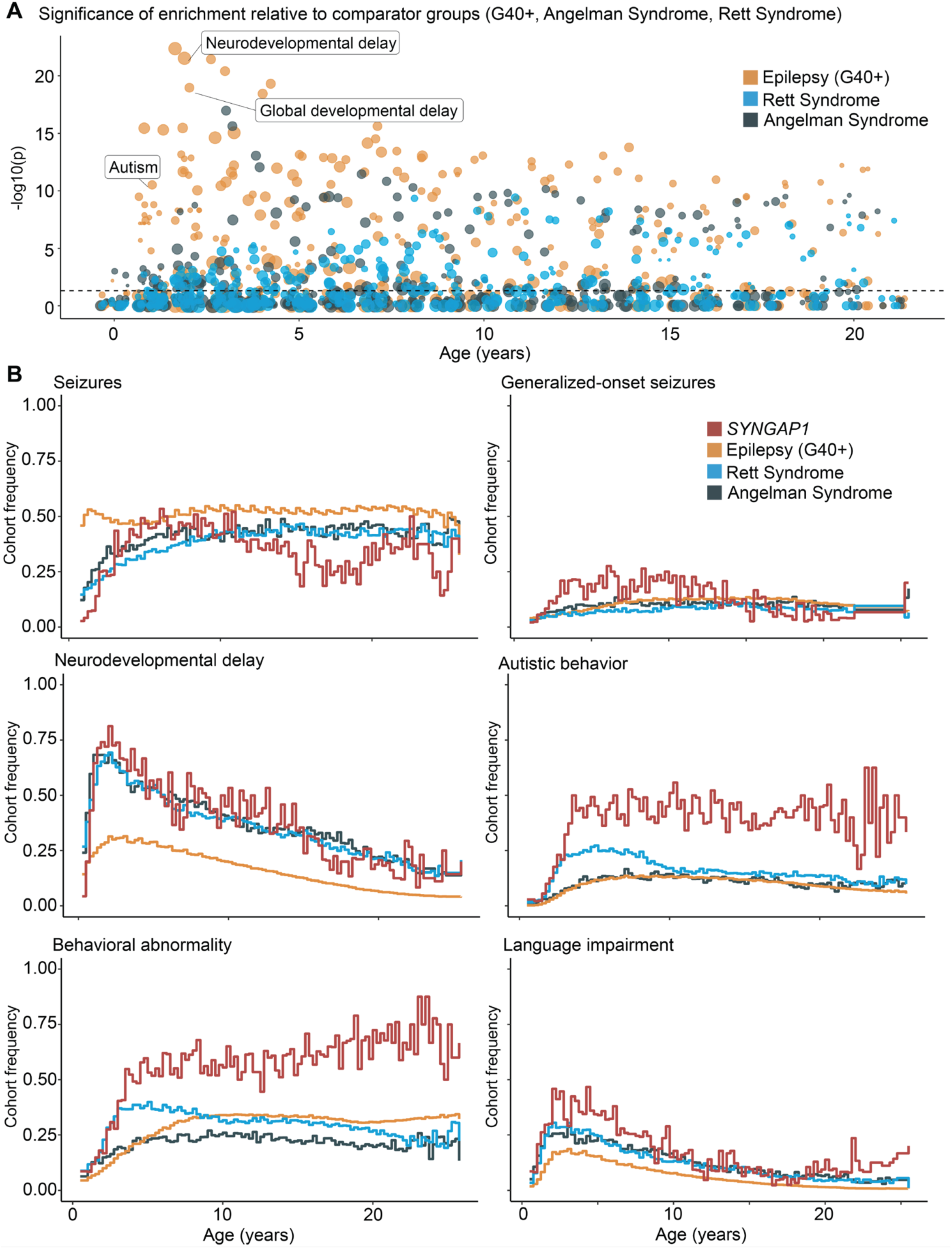
Age-related clinical features in *SYNGAP1* compared to longitudinal histories of other epilepsies and neurodevelopmental disorders. The overall clinical presentation of *SYNGAP1*-RD begins to diverge from comparator groups by the second year of life. (A) The significance of enrichment of key features compared to comparator groups (G40+, Angelman Syndrome, Rett Syndrome) is plotted across the lifespan. (B) The frequencies of selected features across the lifespan are shown for individuals with *SYNGAP1*-RD and comparator groups. When compared to the general epilepsy cohort, we found that individuals with *SYNGAP1* were more likely to have behavioral abnormalities, first significant between 27 and 30 months (OR 3.00, 95% CI 1.50 – 5.68) and persisting throughout the lifespan. Autistic behavior also became prominent between 27 and 30 months (OR 5.71, 95% CI 2.44 – 11.9). Generalized-onset seizures became significantly enriched (OR 4.05, 95% CI 2.02 – 7.59) after 3 years of age.

The majority of individuals with *SYNGAP1-*RD have generalized epilepsy, including generalized-onset seizures in up to 70% (frequency range 66.5%, *n*=107/161 claims data to 69.5%, *n*=91/131 medical records). The frequency of bilateral tonic-clonic seizures ranged from 16.8% (*n*=22/131 in medical records) to 46.6% (*n*=75/161 in claims data), absence seizures in 18.0% (*n*=29/161) to 54.2% (*n*=71/131), atonic seizures in 32.9% (*n*=53/161) to 42.0% (*n*=55/131), and generalized myoclonic-atonic seizures in up to 43.5% (*n*=70/161) with myoclonic seizures captured in *n*=30/131 (22.9%). The median age of seizure onset was 2.8 years (IQR 2.0 – 3.8 years): 6 individuals had infantile onset, 26 in the second year of life, and 40 in the third year of life, and 59 after the third year of life (**Fig 3A**).

**Figure 3.**
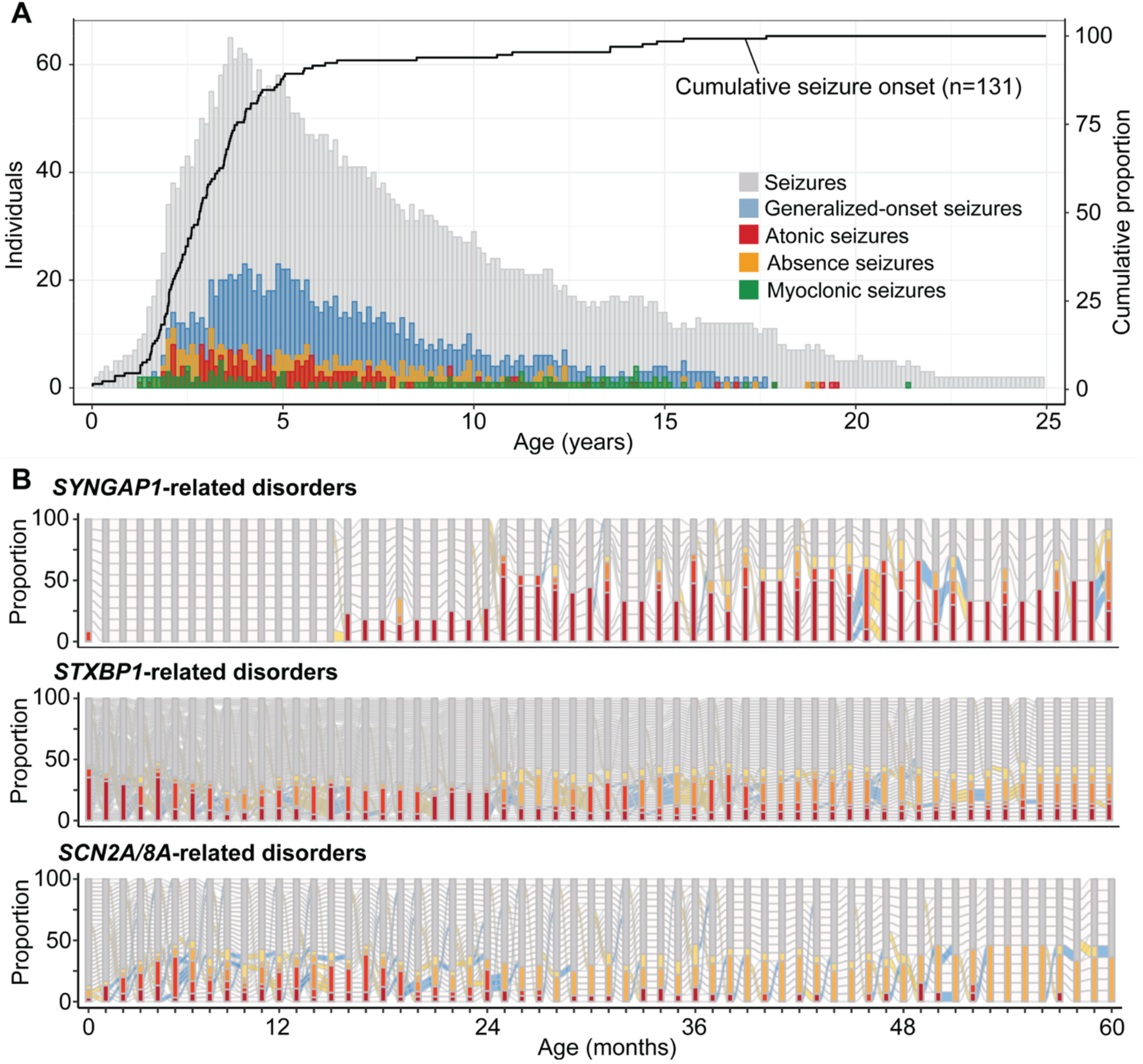
Seizure landscape in *SYNGAP1-*RD differ from other common genetic epilepsies. (A) Generalized-onset seizures occur in up to 70% (frequency range 66.5%, *n*=107/161 claims data to 69.5%, *n*=91/131 medical records). The frequency of bilateral tonic-clonic seizures ranged between 16.8% (*n*=22/131 in medical records) to 46.6% (*n*=75/161 in claims data), absence seizures in 18.0% (n=29/161) to 54.2% (*n*=71/131), atonic seizures in 32.9% (*n*=53/161) to 42.0% (*n*=55/131) and generalized myoclonic-atonic seizures in up to 43.5% (n=70/161). (B) The median age of seizure onset was 2.8 years (IQR 2.0–3.8 years), later than other common genetic epilepsies.

Detailed monthly seizure histories were reconstructed in 13 individuals with *SYNGAP1-*RD across 154 cumulative patient-years. Epilepsy trajectories of *SYNGAP1* are distinct from other genetic epilepsies including *STXBP1*-related disorders and *SCN2A*/*8A*-related disorders (**Fig 3B**). Of individuals with epilepsy, seizures tended to start after the second year of life and were more likely to be refractory with frequent generalized seizures including absence seizures and atonic seizures. When assessing the median seizure frequency, we found that individuals with *SYNGAP1*-RD often had seizure frequencies falling within the highest ELHS/PELHS category (>5 per day) due to the prevalence of absence and myoclonic seizures, which tend to occur at higher frequencies than other seizure types.

### Developmental outcomes and behavioral features are distinct in SYNGAP1-RD

We mapped milestone acquisition in 158 individuals across 1,254 patient-years. The range of individuals achieving specific developmental milestones spanned from less than 23% for the ability to use verbal communication to more than 95% for the ability to communicate using nonverbal language and achieve fine motor abilities (**Fig 5B**). For those individuals achieving each specific milestone, the median age at head control was 3 months (*n*=35, IQR 2 – 6.7 months), rolling over at 5.9 months (*n*=70, IQR 4.0 – 8.6 months), sitting unsupported at 9.8 months (*n*=73, IQR 8.6 months – 1.1 years), walking with or without assistance at 1.7 years (*n*=143, IQR 1.5 – 2.3 years), and using short phrases at 4.2 years (*n*=39, IQR 2.9 – 5.5 years, **Fig 5A**).

**Figure 4.**
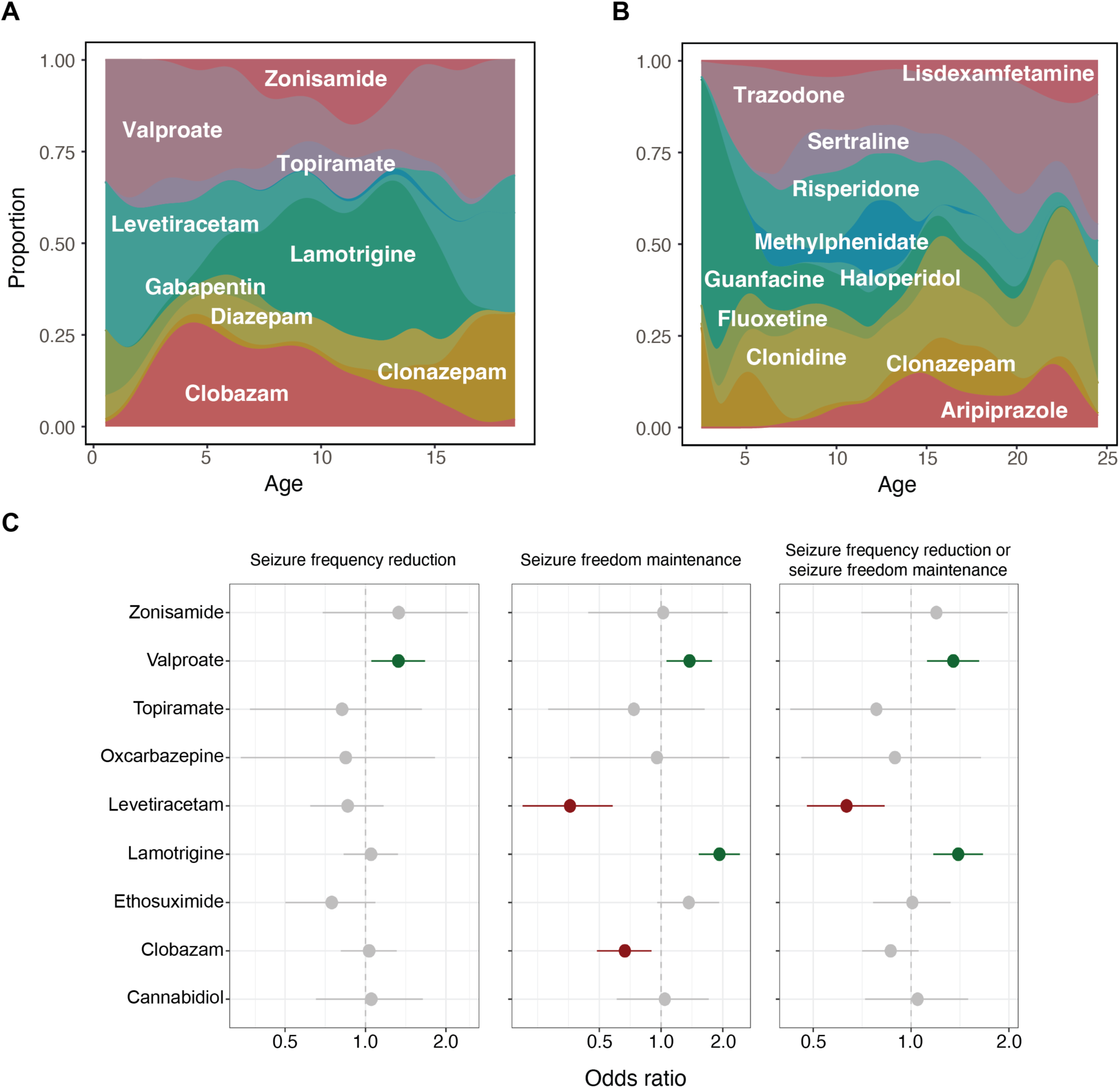
The ASM prescription data reveals *SYNGAP1*-specific treatment approaches. (A) Individuals with *SYNGAP1* are more commonly treated with valproic acid (OR 2.26, 95% CI 1.29 – 3.70) and clobazam (OR 2.58, 95% CI 1.55 – 4.09) for epilepsy and (D) risperidone (OR 5.43, 95% CI 3.47 – 8.18), aripiprazole (OR 3.52, 95% CI 2.05 – 5.69) and guanfacine (OR 2.97, 95% CI 1.76 – 4.75) for behavior, which contrasted from treatment strategies frequently used in the broader epilepsy population. (B) Comparative ASM effectiveness analysis (green – significant association [P < 0.05 with OR > 1], grey – non-significant association, red – nominally significant association [P < 0.05 with OR < 1]). The use of valproate and lamotrigine demonstrated effectiveness in reducing seizure frequencies or maintaining seizure freedom.

**Figure 5.**
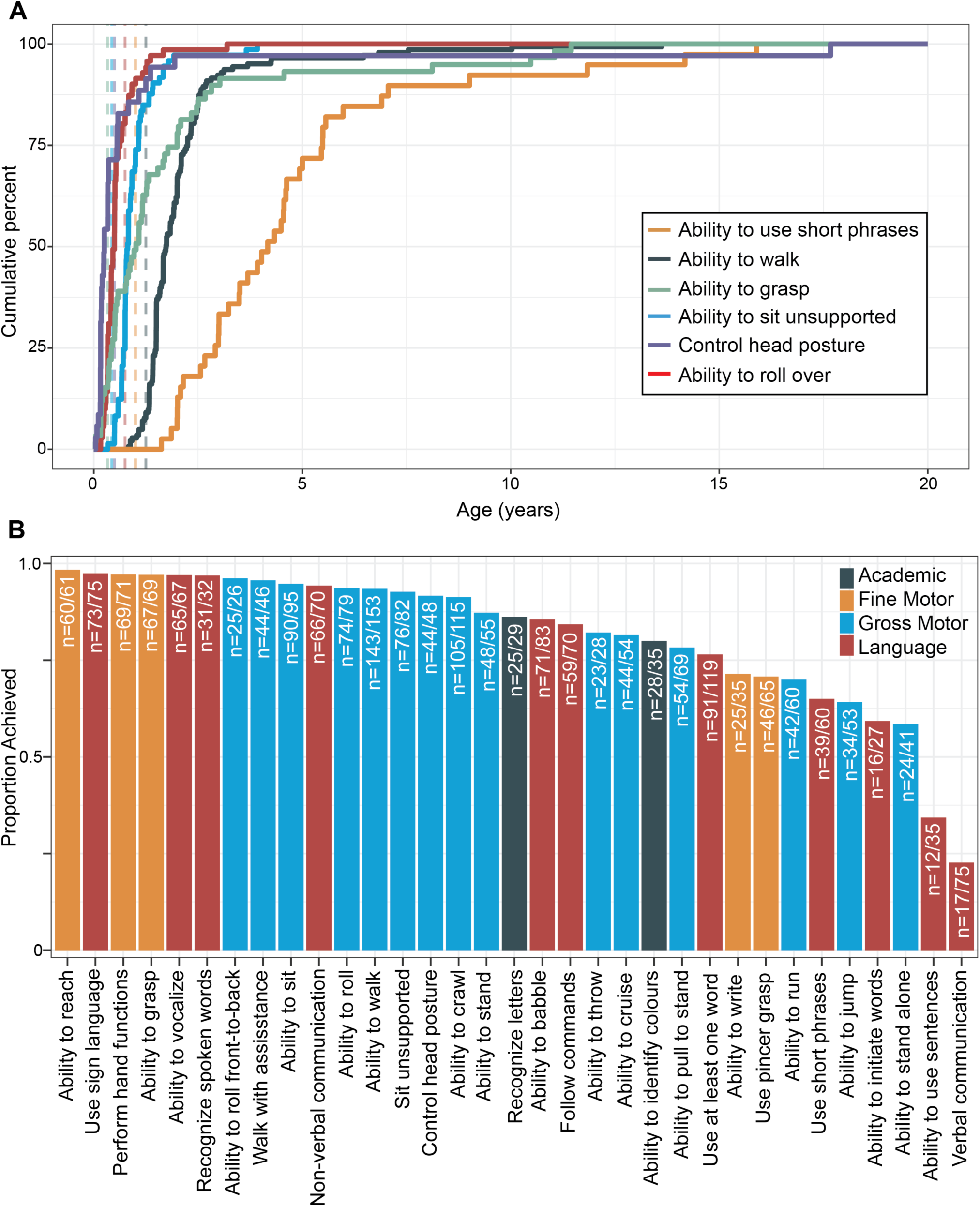
Developmental skill acquisition in 158 individuals across 1,254 patient-years illustrates the unique features of the *SYNGAP1*. (A) The median age at achieving head control was 3 months (*n*=35, IQR 2 – 6.7 months), rolling over at 5.9 months (*n*=70, IQR 4.0 – 8.6 months), sit unsupported at 9.8 months (*n*=73, IQR 8.6 months – 1.1 years), walk with or without assistance at 1.7 years (*n*=143, IQR 1.5 – 2.3 years), and use short phrases at 4.2 years (*n*=39, IQR 2.9 – 5.5 years). The cumulative acquisition of commonly assessed milestones is displayed over the lifespan, for those individuals for which that skill was attained. (B) The range of developmental milestones achieved spanned from 23% in ability to use verbal communication to more than 95% in ability to communicate using nonverbal language and achieve fine motor abilities.

Almost all individuals with *SYNGAP1-*RD have behavioral features (95.7%, *n*=132/138 in medical record cohort), including sleep disturbance in up to 74.6% (*n*=103/138), anxiety-related behavior in up to 23.9% (*n*=33/138), and aggressive behavior in up to 52.2% (*n*=72/138).

### The medication landscape in SYNGAP1 demonstrates etiology-specific treatment strategies

Lastly, we assessed the current treatment landscape of *SYNGAP1-*RD. Leveraging claims data for ASM and behavioral medication prescriptions, we were able to reconstruct the medication histories of 246 individuals across 45 unique medications. For epilepsy therapy (**Fig 4A** & **Fig S2**), we found that valproic acid (OR 2.26, 95% CI 1.29 – 3.70) and clobazam (OR 2.58, 95% CI 1.55 – 4.09) were more frequently used in individuals with *SYNGAP1* in comparison to the broader cohort of individuals with epilepsy (G40+). The medication landscape for behavioral features in *SYNGAP1-*RD (**Fig 4B** & **Fig S2**) demonstrated that risperidone (OR 5.43, 95% CI 3.47 – 8.18), aripiprazole (OR 3.52, 95% CI 2.05 – 5.69) and guanfacine (OR 2.97, 95% CI 1.76 – 4.75) were more commonly used for managing behavioral symptoms.

In terms of the ASM effectiveness (**Fig 4C**), we found that valproate (*n* = 43, *P* = 0.016, OR 1.33, CI 1.05-1.67) had a significant association with reductions in seizure frequencies as well as with maintaining seizure freedom (*n* = 43, *P* = 0.013, OR 1.38, CI 1.06-1.77). Lamotrigine (*n* = 35, *P* = 2.91 x 10^−8^, OR 1.93, CI 1.53-2.42) also showed significant associations with maintaining seizure freedom. In contrast, levetiracetam (*n* = 44, *P* = 2.23 x 10^−6^, OR 0.36, CI 0.21-0.58) and clobazam (*n* = 47, *P* = 6.43 x 10^−3^, OR 0.67, CI 0.49-0.90) were associated with the lack of seizure freedom.

## DISCUSSION

Disease-causing variants in *SYNGAP1* result in a recognizable neurodevelopmental disorder characterized by generalized epilepsy, developmental delay, intellectual disability, and behavioral features. However, there is considerable variability in disease course, severity, and functional outcome. As novel precision medicine therapeutics are under development for rare genetic epilepsies, including *SYNGAP1*, complete characterization of the phenotypic landscape and developmental outcomes of the condition is critical for clinical trial readiness.^13,14,29–31^

While well-designed natural history studies with validated outcome measures are the ideal precursor to clinical trial design, such studies are expensive, time-consuming, and have difficulties achieving enrollment targets, especially in rare disease. ^32^ The current study lays the groundwork for future natural history studies by leveraging real-world data across multiple healthcare resources to delineate the longitudinal epilepsy and phenotypic landscape of *SYNGAP1*-RD. The use of EMR and claims data also allows for the comparison of longitudinal phenotypes in *SYNGAP1*-RD to other similar syndromic comparator groups, which allows us to identify early phenotypic fingerprints of these disorders.

### Phenotypic signatures enriched in SYNGAP1-RD

We capitalized on three unique real-world datasets to delineate both the overall burden of clinical features as well as longitudinal symptom trajectories. Our analyses show a significant enrichment of autism and behavioral abnormalities starting at 27 months, representing the earliest distinguishing features of this disorder, when compared to a general epilepsy cohort. Generalized seizures were also a key feature, but only became significantly enriched after 3 years of age. Furthermore, when analyzed relative to two similar syndromic comparator groups—Angelman and Rett Syndromes—the relative enrichment of both autism and behavioral features in *SYNGAP1-*RD held true.

### Epilepsy onset and trajectories

Seizure histories in *SYNGAP1-*RD are unique among the genetic epilepsies, with relatively later onset (median 2.8 years) and a tendency towards persistent frequent seizures throughout childhood. Most common seizure types were generalized, including absence, atonic and myoclonic. Of note, bilateral tonic-clonic seizures were present in 47% of those with seizures reported in claims data, while only in 17% of those in the EMR datasets. Two possible explanations are that (1) more severe seizures are more likely to be coded in claims data, thus inflating the proportion of significant motor seizures while underrepresenting milder seizure types such as absence, and (2) that non-neurologists often document all seizures with motor signs as “generalized tonic-clonic seizures (GTCs),” introducing a systemic error to the claims dataset.

In our study, we identified an overall lower rate of epilepsy compared to previously published cohort studies.^4,7^ This is likely due to two factors—the continual improvement in access to genetic testing has led to earlier and broader testing, including those individuals without seizures and milder phenotypes, and the natural limitations of claims and EMR-based datasets. Historically, only the most severely affected individuals were offered genetic testing, but as it becomes more ubiquitous, we expect to see a widening of the phenotypic spectrum, encompassing more mild cases over time.

### Developmental outcomes and behavioral features

Development in *SYNGAP1-*RD follows a distinct trajectory, with global delays in milestone achievement, however, language acquisition is reliably the most severely affected. In our clinical experience, behavioral concerns are often the most significant issue raised by caregivers of individuals with *SYNGAP1-*RD, which has previously been demonstrated by smaller case-series^33^ and recapitulated here in our large, retrospective, real-world datasets. That behavioral medications are so frequently prescribed also underscores the impact of the behavioral symptoms in this disorder.

### Current state of treatment strategies

While targeted therapeutics are currently under development,^13–15^ medical treatment of *SYNGAP1*-related epilepsy and behavior follows typical treatment practices for other generalized epilepsies and behavioral disorders. Seizures are most commonly treated with clobazam and valproic acid, while behavioral medications are very frequently prescribed, with the most common choices being risperidone, guanfacine, and aripiprazole. Comparative effectiveness analysis confirmed the efficacy of valproic acid and lamotrigine for reducing seizure frequency and maintaining seizure freedom. This confirms our experience in clinical practice and is intuitive as these are both effective medications for generalized seizures. However, neither clobazam nor levetiracetam were shown to be effective in this cohort. This may be related to a lack of power to detect a significant reduction in seizures, but may also be due to the clinical characteristics of these individuals. In particular, levetiracetam is often prescribed for eyelid myoclonia, so our results may be reflective of the inherent high frequency and treatment-resistance of this seizure type. Additionally, one unique feature about the claims dataset is the relatively low capture of cannabidiol prescriptions; this appears to be specific to the data vendor used in these analyses and may be due to specialty pharmacy blocking. While the current treatment strategies are reactionary—treating symptoms as the arise—identifying individuals with *SYNGAP1-*RD early will be critical once the clinical trials for targeted therapies are available.

### Limitations

While the primary mechanism of disease in *SYNGAP1-*RD is haploinsufficiency, prior studies have suggested possible genotype-phenotype correlations, including the lower prevalence of epilepsy in individuals with variants in the SH3-binding motif,^12^ the relatively milder presentations in individuals with loss of function variants in exons 1-4 compared to exons 5-19,^7,12^ and the relative pharmacosensitivity of the epilepsy in individuals with variants in exons 4 and 5.^6^ The variant spectrum from medical record data included 134 PTVs, 21 missense variants, and two complex or in-frame indels. Given the limitations of claims data, we were unable to link the phenotypic information to a genetic testing report. However, techniques for tokenization of the data are under development which will make linking claims data to genetic data feasible, greatly enhancing the utility of these large real-world datasets.^34^ This would also allow for greater harmonization across diverse data sets, ensuring individuals contained in each dataset could be identified and linked, enhancing the quality of their representation.

Due to relatively large sample sizes, claims data can provide useful insights into longitudinal disease trajectories; however, to that the data is sparse and subject to unique challenges.^35,36^ These datasets are prone to errors in medical coding, and any errors are often perpetuated due to the tendency to “copy forward” diagnosis codes from prior encounters. Furthermore, while we are relatively confident that clinical features coded at high frequencies are accurate, the absence of coding of a feature or symptom cannot be taken to mean that feature was not present, as not all clinical symptoms are captured in encounter diagnosis codes. This likely accounts for the differences in frequencies of given clinical features between the claims dataset and the combined electronic medical record dataset—for example, epilepsy was reported in 82.9% of cases obtained through medical records, but only 65.4% of those obtained through insurance claims data.

## Conclusion

Taken together, we surveyed the clinical landscape of *SYNGAP1-*RD and contrasted the phenotypic signature and current state of treatment strategies with other epilepsies and neurodevelopmental disorders, including Angelman Syndrome and Rett Syndrome. This analysis underscored the relatively later onset of a generalized epilepsy with very prominent behavioral features, autism, and significant impairements in expressive language. The integration and analysis of real-world data from various healthcare databases enabled us to leverage varying scopes of data granularity and complexity, demonstrating the utility of large-scale, computational frameworks to understand the prevalence of clinical features and outline the longitudinal landscape of rare diseases. Delineation of disease-specific trajectories in *SYNGAP1-*RD will remain critical for prospective natural history studies, clinical trial-readiness, and future precision medicine advances.

## Data Availability

De-identified aggregate data will be available upon written request to the corresponding author. The raw data that support the analyses presented here contain sensitive and protected health information for participants and are therefore not openly available.

## ACKNOWLEDGMENTS

This study was supported by the Center for Epilepsy and Neurodevelopmental Disorders (ENDD), the National Institute for Neurological Disorders and Stroke (R01 NS127830-01A1, R01 NS131512-01 and K02 NS112600 to IH), the American Epilepsy Society (AES), Pediatric Epilepsy Research Foundations (PERF) & SynGAP Research Fund (SRF) through a Research Training Fellowship for Clinicians (JLM), and the American Academy of Neurology (AAN), AES, the Epilepsy Foundation, & the American Brain Foundation (ABF) through the Susan Spencer Award (JLM).

## AUTHOR CONTRIBUTIONS

1. conception and design of the study (JLM, IH, JX, RS)
2. acquisition and analysis of data (JLM, JHM, JX, SC, JT, CC, DK, AR, SMR, RS, MJB, EB, NF, JMG, KAH)
3. drafting a significant portion of the manuscript or figures (JLM, JHM, JX)

