## Supplemental Figures for "Clinical signatures of *SYNGAP1-*related disorders through data integration"

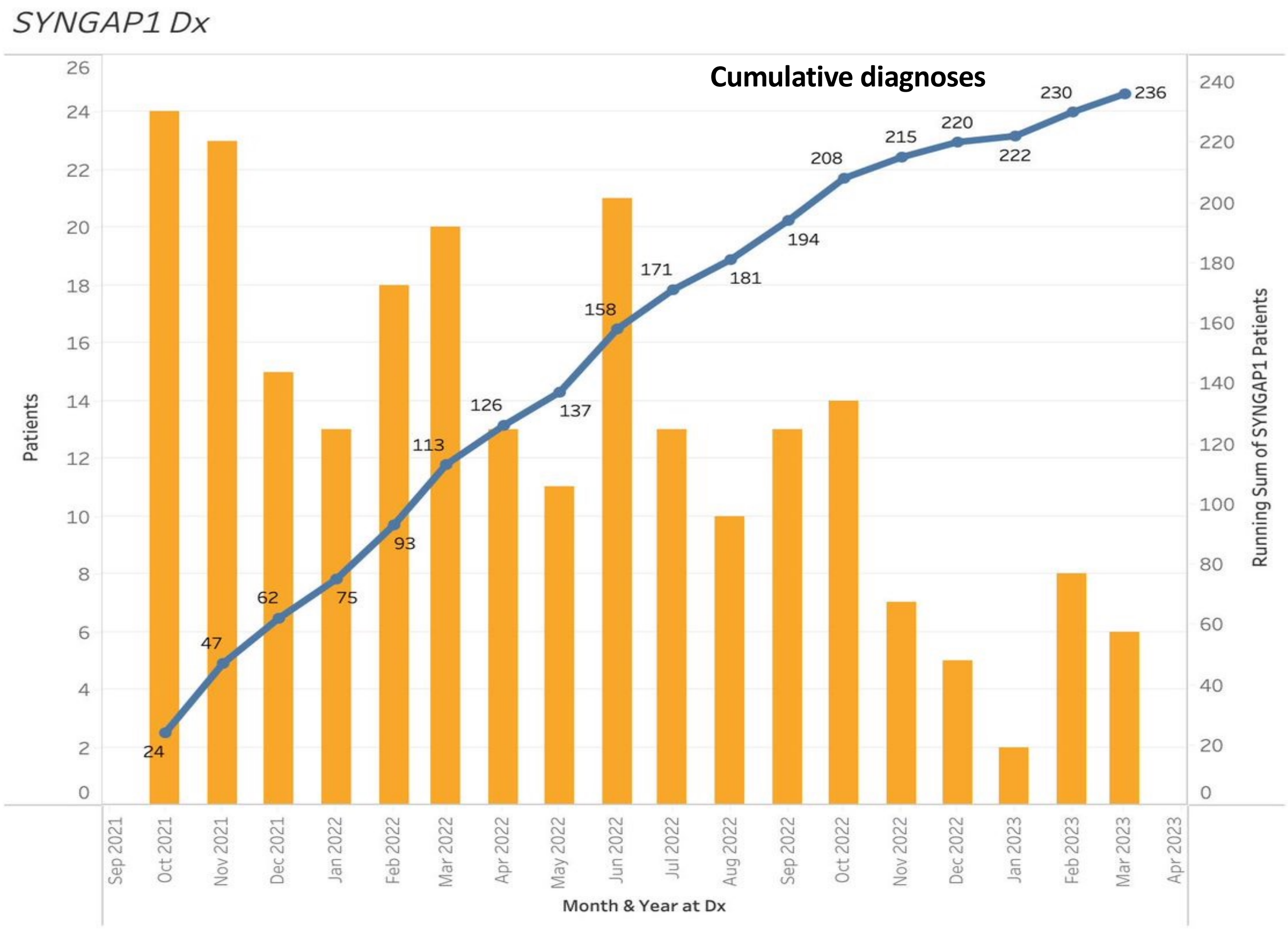


**Supplementary Figure 1.**

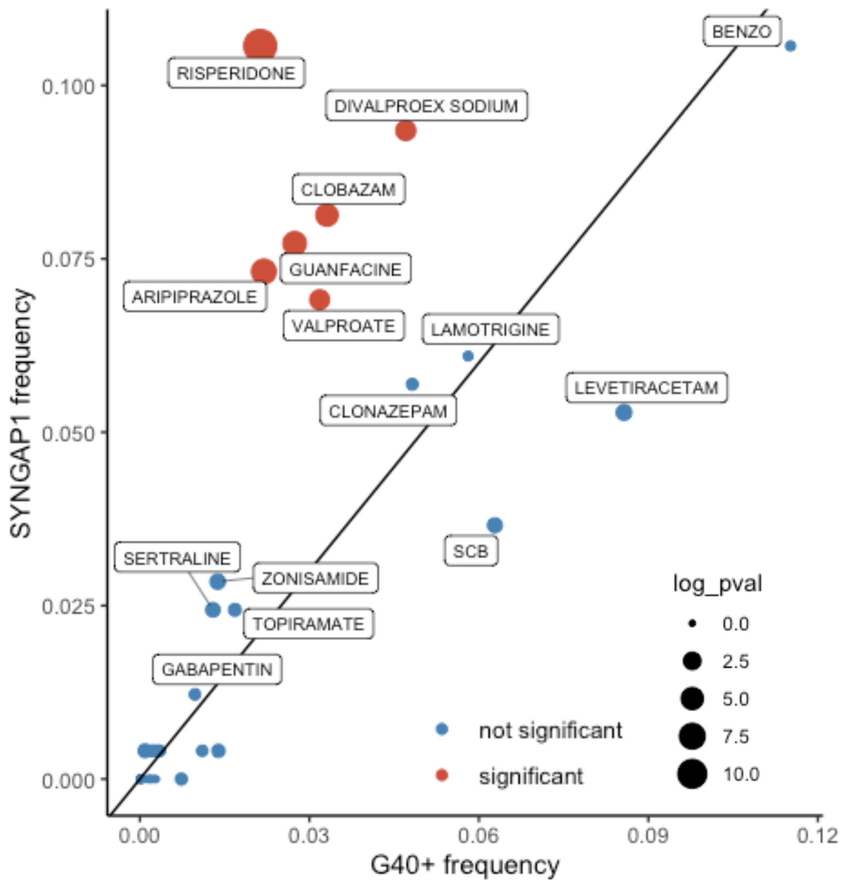


**Supplementary Figure 2. More commonly prescribed medications in *SYNGAP1*-related disorders compared to the broader epilepsy cohort.**
